# Exploring the Pitfalls of Large Language Models: Inconsistency and Inaccuracy in Answering Pathology Board Examination-Style Questions

**DOI:** 10.1101/2023.08.03.23293401

**Authors:** Shunsuke Koga

**Affiliations:** Department of Pathology and Laboratory Medicine, Hospital of the University of Pennsylvania, Philadelphia, Pennsylvania, USA

**Keywords:** artificial intelligence, large language models, pathology, ChatGPT, Google Bard, inconsistency, comparative study, application in medicine

## Abstract

In the rapidly advancing field of artificial intelligence, large language models (LLMs) such as ChatGPT and Google Bard are making significant progress, with applications extending across various fields, including medicine. This study explores their potential utility and pitfalls by assessing the performance of these LLMs in answering 150 multiple-choice questions, encompassing 15 subspecialties in pathology, sourced from the PathologyOutlines.com Question Bank, a resource for pathology examination preparation. Overall, ChatGPT outperformed Google Bard, scoring 122 out of 150, while Google Bard achieved a score of 70. Additionally, we explored the consistency of these LLMs by applying a test-retest approach over a two-week interval. ChatGPT showed a consistency rate of 85%, while Google Bard exhibited a consistency rate of 61%. In-depth analysis of incorrect responses identified potential factual inaccuracies and interpretive errors. While LLMs have potential to enhance medical education and assist clinical decision-making, their current limitations underscore the need for continued development and the critical role of human expertise in the application of such models.

## Introduction

Over the past decade, artificial intelligence (AI) has made significant progress, particularly in the development of large language models (LLMs). These models use extensive text data for training, enabling them to generate human-like text, understand context, respond to queries, and facilitate language translation ^1^.

ChatGPT, including its advanced versions GPT-3.5 and GPT-4, is an LLM developed by OpenAI that has been extensively used in a range of applications, such as writing assistance and programming support. Another notable LLM is Google Bard developed by Google. Importantly, Google Bard has the unique ability to access real-world, current information through Google Search. This capability renders it especially beneficial for answering queries that require the latest data.

LLMs, including ChatGPT, have demonstrated their potential in medical education ^2, 3^. ChatGPT not only exhibited promising performance in the United States Medical Licensing Exam ^4^, but also in equivalent medical exams in Japan and Italy ^5, 6^. Current research is examining how LLMs perform in particular medical disciplines. For instance, GPT-4 outperformed other LLMs like GPT-3.5 and Google Bard in a neurosurgery oral boards examination^7^. In another study comparing ChatGPT and Google Bard’s responses to lung cancer inquiries, both models showed accuracy, though not without flaws ^8^.

While AI and machine learning techniques, particularly image analysis ^9-11^, have been explored in pathology, the evaluation of LLMs has not been extensively investigated ^12, 13^. This study seeks to address this gap by assessing the performance of ChatGPT and Google Bard within the field of pathology. The focus is on evaluating and comparing their accuracy and consistency in answering board examination-style questions in pathology.

## Methods

### Question Selection and Evaluation

The study compared the performance of two LLMs, ChatGPT (GPT-4) and Google Bard, using questions from the PathologyOutlines.com Question Bank (https://www.pathologyoutlines.com/review-questions), a resource for pathology examination preparation.

The question bank had 3365 questions across pathology subspecialties, but for this study, we selected 150 multiple-choice questions, with 10 from each of 15 subspecialties, to ensure a balanced dataset: Autopsy & forensics, Bone, joints & soft tissues, Breast, Dermatopathology, Gastrointestinal & liver, Genitourinary & adrenal, Gynecological, Head & Neck, Hematopathology, Informatics & Digital Pathology, Medical renal, Neuropathology, Stains & CD markers/Immunohistochemistry, Thoracic, and Clinical Pathology. Each question was presented in a single best answer, multiple-choice format. Both LLMs were presented with the same set of questions. No additional context or hints were provided to the models apart from the questions themselves, to simulate real-world application. Questions containing images were excluded from the question bank as ChatGPT is not capable of processing image data.

To evaluate the consistency of both LLMs, a test-retest approach was implemented. The same set of 150 questions was posed to the models on two separate occasions, with a two-week interval. By comparing the answers on both performances, we aimed to assess the consistency in their responses.

### Statistical analyses

Statistical analyses were performed using R version 4.3.1 (R Foundation for Statistical Computing, Vienna, Austria). A Chi-square test was conducted to compare the performance of ChatGPT and Google Bard. Statistical significance was set at p < 0.05.

## Results

### Overall test scores

The performance of the two LLMs, ChatGPT and Google Bard, was evaluated across 15 subspecialties in pathology. Overall, ChatGPT significantly outperformed Google Bard across all subspecialties; ChatGPT achieved a total score of 122 out of 150, compared to Google Bard’s score of 70 (p < .001). Detailed performance outcomes of each LLM across all subspecialties are presented in **Table 1**.

**Table 1:** Performance scores of ChatGPT and Google Bard across pathology subspecialties.

| **Subspeciality** | **ChatGPT** | **Google Bard** |
| --- | --- | --- |
| Autopsy & Forensics | 9 | 6 |
| Bone, Joints & Soft Tissues | 7 | 3 |
| Breast | 7 | 3 |
| Dermatopathology | 8 | 5 |
| Gastrointestinal & Liver | 9 | 6 |
| Genitourinary & Adrenal | 6 | 4 |
| Gynecological | 9 | 7 |
| Head & Neck | 9 | 3 |
| Hematopathology | 9 | 5 |
| Informatics & Digital Pathology | 9 | 7 |
| Medical Renal | 9 | 3 |
| Neuropathology | 8 | 5 |
| Stains & CD markers/Immunohistochemistry | 8 | 3 |
| Thoracic | 8 | 5 |
| Clinical Pathology | 7 | 5 |
| **Total** | **122** | **70** |

### Assessment of Consistency

In the assessment of consistency of both LLMs, test scores were largely consistent between the first and second sessions. The scores of ChatGPT were 122 and 126 out of 150 in the first and second tests, respectively, while Google Bard scored 70 and 69 in the same tests. Despite the relative stability in test scores, a detailed inspection revealed significant changes; identical answers in both sessions were present in only 85% (127/150) of ChatGPT’s responses and a lower 61% (92/150) of Google Bard’s (**Table 2**). ChatGPT initially provided 28 incorrect answers. In the retest, it corrected 11 of these but also altered 7 correct answers to incorrect ones. Among the initial errors, ChatGPT repeated 5. Google Bard exhibited a similar but more pronounced pattern. Starting with 80 incorrect responses, it corrected 19 in the retest, made new errors in 20 previously correct answers, and repeated 19 of its original mistakes in the retest.

**Table 2:** Consistency of Model Responses.

| **Outcome** | **ChatGPT** | **Google Bard** |
| --- | --- | --- |
| No change in response | 127 (85%) | 92 (61%) |
| Correct to incorrect response | 7 (5%) | 20 (13%) |
| Incorrect to another incorrect response | 5 (3%) | 19 (13%) |
| Incorrect to correct response | 11 (7%) | 19 (13%) |

### Evaluation of Incorrect Responses

We identified incorrect answers from both LLMs in our study. One example was a question about Lynch syndrome: “Lynch syndrome usually arises from a germline mutation in a gene coding for a mismatch repair protein. A germline mutation in which of the following genes could also cause Lynch syndrome?” with options A. BRAF, B. CDH1, C. EPCAM, D. MUTYH. The correct answer was C. EPCAM ^14^. However, Google Bard incorrectly chose option D. MUTYH and justified its answer by associating MUTYH mutations with Lynch syndrome, which is a factual inaccuracy as MUTYH mutations cause a different type of hereditary colorectal cancer known as MUTYH-associated polyposis ^15^. In contrast, ChatGPT correctly selected the answer as C. EPCAM.

Another example was a question about nemaline myopathy: “In which gene are de novo mutations most commonly associated with nemaline myopathy?” with options A. NEB, B. KLHL40, C. TPM3, D. ACTA1, and E. TNNT1. The correct answer was D. ACTA1, but both models selected A. NEB. Specifically, ChatGPT justified its answer by stating that NEB is the most commonly involved gene in cases of nemaline myopathy, while Google Bard indicated that de novo mutations in the NEB gene are the most common cause of this condition. These responses exhibit factual and interpretative inaccuracies, as NEB mutations, while common in nemaline myopathy, are typically inherited in an autosomal recessive manner, not de novo ^16^.

## Discussion

Our study further underscores the strengths and weaknesses of LLMs in medicine. While ChatGPT consistently surpassed Google Bard in accuracy and consistency, neither model answered all questions correctly, suggesting knowledge or comprehension gaps. Additionally, retesting after two weeks revealed inconsistencies in both LLMs’ responses to the same questions, highlighting potential reliability issues ^17^.

In assessing LLMs’ comprehension of medical queries, our study identified two error types. Firstly, Google Bard displayed factual inaccuracies, incorrectly linking MUTYH to Lynch syndrome despite vast data access. Secondly, ChatGPT exhibited interpretation errors. While answering a question about “de novo” mutations in nemaline myopathy, it correctly identified NEB as a common cause but overlooked the specific “de novo” context, highlighting LLMs’ potential for nuanced misunderstandings.

Another important consideration in the application of LLMs in medical fields is their consistency or reliability, defined as the models’ ability to provide the same answer to identical prompts when asked on different occasions. Our assessment of consistency revealed a suboptimal consistency rate for both LLMs (i.e., 85% in ChatGPT and 61% in Google Bard), which are consistent with the results of another study that evaluated ChatGPT’s responses to surgical case questions ^18^. Such inconsistencies underline the current limitations of LLMs and highlights the necessity for further development and refinement to improve their consistency for effective use in the medical field.

There are some limitations of our study. Firstly, there was no direct comparison with human performance. While our results shed light on the capabilities of LLMs in answering complex medical questions, understanding how their performance compares directly to medical students or professionals remains crucial. Additionally, our focus was largely on pathology questions in English language. To generalize our findings, future studies should encompass different medical specialties and languages. Lastly, the challenge of incorporating images into our evaluation also presents a significant limitation.

In conclusion, our study indicates that LLMs have the potential to assist in clinical decision-making in the future. Both models demonstrated inconsistencies and inaccuracies, emphasizing the need for their further development and rigorous validation. While the potential of these AI models is promising, human oversight and expertise remain crucial in the medical field.

## Data Availability

All data produced in the present study are available upon reasonable request to the author

## Abbreviations

AI: Artificial Intelligence
LLMs: Large Language Models

## Acknowledgement

This manuscript was edited and proofread by ChatGPT (GPT-4, OpenAI), and the author verified the final content.

## Author Contributions

Conception and design of the study: SK

Acquisition and analysis of data: SK

Drafting the manuscript: SK

